# Systematic Review of Knowledge, Attitudes, and Practices of Dairy Farmers and Consumers Towards Bovine Tuberculosis in Low- and Middle-Income Countries

**DOI:** 10.1101/2024.04.19.24306060

**Authors:** Abigail Van Der Zwan, Patricia T. Campbell, Nancy Shi, Nikita De Bortoli, Juan Pablo Villanueva-Cabezas

## Abstract

**Background:** Bovine Tuberculosis (bTB), caused by *Mycobacterium bovis*, is a neglected zoonotic disease primarily associated with cattle. The incidence of bTB is highest in low-income settings with high cattle density and unpasteurised dairy consumption. Smallholder dairy farming has been steadily growing in low- and middle-income countries (LMICs) with limited professional support for adequate bTB surveillance and risk mitigation. Several studies have explored the knowledge, attitudes, and practices (KAP) of milk value chain’s stakeholders towards bTB in LMICs, but this evidence has not been collated and synthesised.

**Methodology:** We conducted a systematic review to determine what is known, believed, and done in relation to bTB among dairy producers and consumers in LMICs. We performed a systematic search of studies in OVID Medline, Scopus and CABI on 11 September 2023. KAP data were summarised using narrative synthesis and forest plots.

**Results:** We retrieved 2763 articles, retaining 51 for the review. Only studies from Africa (n=38) and Asia (n=13) met the eligibility criteria. Most populations reported awareness of human tuberculosis and knew it could be treated, but there was limited awareness of bTB and its zoonotic potential. Knowledge of bTB transmission routes and bTB mitigation varied and risky practices were also variable across populations. Inconsistencies in study design and survey tools suggest some results may have mid- to high risk of bias.

**Conclusions:** Awareness of bTB is surprisingly low among African and Asian populations with high bTB exposure risk, possibly due to the long-standing divide between animal and human health messages that has obscured the One Health implications of bTB. Addressing bTB in LMICs requires a structural One Health approach and standard KAP survey tools to adequately explore the socio-cultural, political, and economic processes and drivers favouring bTB spread and persistence.

## 1 Introduction

Every year, tuberculosis causes 10 million new infections and 1.5 million deaths worldwide (1). Human tuberculosis is caused by *Mycobacterium tuberculosis* (2) but up to 10% of human cases are caused by *Mycobacterium bovis (M. bovis),* the zoonotic bacteria that causes bovine tuberculosis (bTB) (3). Cattle are the primary reservoir of *M. bovis* and the most significant source of human bTB through consumption of unpasteurised milk, dairy products, raw meat, and inhalation of aerosols (4, 5). While the adverse public health effects of bTB in high-income nations have been mitigated through surveillance, prevention and control (6), bTB remains a global neglected zoonosis that receives insufficient research, policy and public health attention in relation to its health and economic impacts, especially in low- and middle-income settings (7).

The incidence of bTB is prominent in settings with a high density of cattle and people living in poverty who consume unpasteurised dairy products (8). In the last few decades, smallholder dairy farming has steadily grown in less affluent countries with limited professional support for adequate disease surveillance and risk mitigation (9, 10), and a lack of the technologies and systems to perform quality control required in formal milk value chains (11). Although smallholder dairy farming has had positive flow-on livelihood effects for approximately one billion people living in poverty (8), the lack of DIVA (differentiation of infected from vaccinated animals) vaccines (12) and diagnostic methods to quickly and accurately (i.e., high positive and negative predictive values) screen animals, milk, and dairy (13) has translated into highly variable bTB prevalence within herds, and across systems and nations. Uncontrolled bTB increases the exposure risk of producers and dairy consumers; international estimates suggest the prevalence of *M. bovis* in bulk tank milk (i.e., before pasteurisation) could be as high as 21% (14), with variable survival of the bacteria associated with different dairy product manufacturing (15, 16).

The roadmap for zoonotic tuberculosis (16) aligns with the World Health Organization’s (WHO) strategy to end the tuberculosis epidemic by 2030 (17), advocating for the diagnosis and treatment of every person with tuberculosis and calling for a paradigm shift that puts people at risk of zoonotic tuberculosis as deserving greater attention. As most populations at risk of bTB live in low- and middle-income countries (LMICs) where the disease is uncontrolled and standard test-and-slaughter programs are socio-economically unfeasible (12), gaining insight into social behaviour towards bTB is crucial. According to WHO, a well-designed and rigorously conducted Knowledge, Attitudes and Practices (KAP) survey produces informative, insightful, and broadly useful data in the planning of evidence-based advocacy, communication, and social mobilisation (18). These activities in turn lead to sustainable social and behavioural change towards tuberculosis (18) needed to reduce exposure to bTB and increase diagnosis and treatment uptake. Over the years, several studies have explored the KAP of milk value chain’s stakeholders towards bTB in LMICs. However, there is yet to be a systematic review that collates and synthesises the evidence generated. Thus, this study aims to collate, summarise and report existing KAP of dairy producers and consumers towards bTB in low- and middle-income countries.

## 2 Methods

This systematic review was registered on PROSPERO on 10 July 2023 (CRD42023438664) and is reported according to the PRISMA 2020 guidelines (19).

### 2.1 Search Strategy

A search of three databases (OVID Medline, Scopus and CABI) was conducted on 11 September 2023. The search terms included “bovine tuberculosis”, “zoonotic tuberculosis”, “mycobacterium bovis”, “M. bovis”, “knowledge”, “attitude”, “perception”, and “practice”. The detailed search strategies for each database can be found in Supplementary Section S.1.

### 2.2 Study Screening

The records retrieved were imported into Covidence (20) for de-duplication, title, abstract and full-text assessment, performed by three independent reviewers (AV, NB and NS). Conflicts were resolved by a fourth independent reviewer (JPV-C).

### 2.3 Eligibility Criteria

The following eligibility criteria were applied: a) The study involved a KAP survey explicitly addressing knowledge, attitudes or practices towards bTB; b) The study participants were dairy farmers or consumers in the milk value chain of any domesticated dairy animal; and c) The study was conducted in a low, lower-middle, or upper-middle income country or countries as per 2023 World Bank classifications (21); d) Full-text journal articles published in any language and any year. The following exclusion criteria were applied: a) Study participants were stakeholders in the farming industry but not directly involved in the milk value chain (e.g., meat cattle farmers or abattoir workers); b) Study participants worked in fields with implied expert knowledge about bTB, such as veterinarians or scientists; and c) The study did not present quantitative estimates of KAP towards bTB.

### 2.4 Data Extraction

The following items were extracted into a Microsoft Excel spreadsheet: author(s), year published, country, sampling method, survey technique, population type, education (% secondary educated and % tertiary educated), gender, and age range of the study participants. Where the sample population was poorly described, information was extracted to contextualise its position in the milk value chain and determine eligibility. Numerators, denominators, and their corresponding fractions were extracted from each study. In cases where studies did not report all three values, missing values were calculated if sufficient information was provided. Finally, a detailed list of KAP questions extracted can be found in Supplementary Section S.2.

### 2.5 Data Synthesis

A narrative synthesis was used to summarise KAP data which were consistently explored across the studies and were relevant to our research question. Key KAP themes were presented in forest plots generated using R Studio Version 4.2.3 (22–24).

### 2.6 Risk of Bias Assessment

The risk-of-bias assessment used the JBI tool for Studies of Prevalence (25), which comprised nine questions (Supplementary Section S.2). To assess overall risk of bias, we assigned one point to ‘Yes’ responses for questions 1, 2, 3, 4, and 9 and 0.5 points to ‘Yes’ responses for questions 5, 6, 7, and 8, deemed less relevant to KAP studies. A risk-of-bias score was calculated as the average of the points assigned and used to categorise studies as having a high (<0.5), moderate (0.5 – 0.75), or low risk of bias (>0.75).

## 3 Results

### 3.1 Study Selection

The literature search yielded 2,763 studies. After removing duplicates, 1,672 studies were screened at the title and abstract level, and 71 were selected for full-text assessment. Of these, 51 studies met the inclusion criteria **(Figure 1)**.

**Figure 1.**
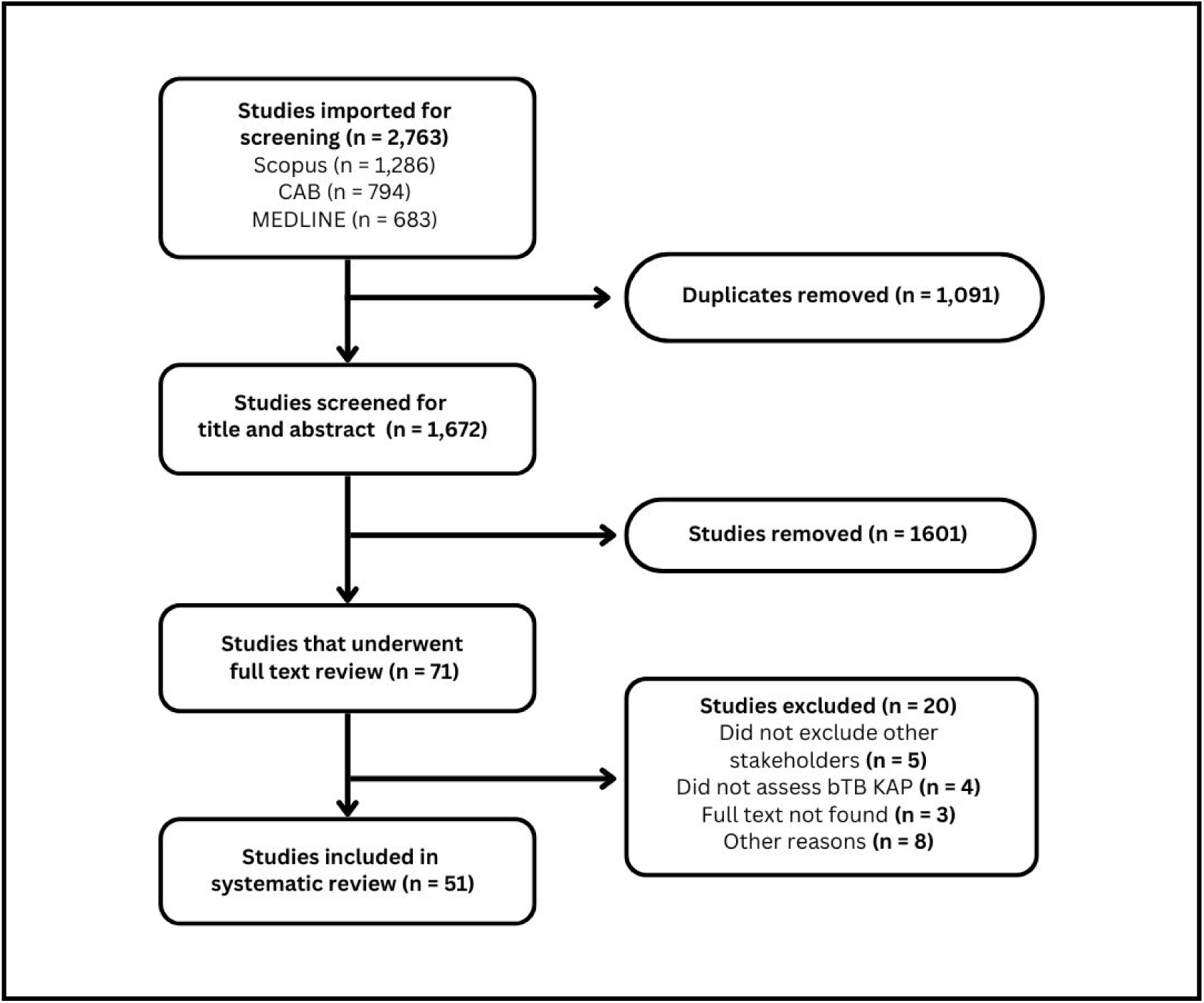
PRISMA flow diagram of literature search and study selection for inclusion in the systematic review.

### 3.2 Study Characteristics

Fifty-one studies published between 2000 and 2023 were included. Most studies (n=38) were conducted across Africa: Ethiopia, Nigeria, Ghana, Niger, Zambia, Zimbabwe, Kenya, Tanzania, Cameroon, South Africa, and Malawi. The rest (n=13) were conducted across Asia: Nepal, India, Bangladesh, Pakistan, and Iraq. In 49 studies, the sample size ranged between 15 and 859, and two studies did not report the number of participants (26, 27). Among the African studies, 30 recruited dairy farming populations — including cattle owners, herdsmen, dairy farm workers or owners, and livestock keepers — and eight recruited pastoralist communities (seven of cattle and one of camel). Three African studies recruited dairy consumers, including the general population, high-school students, and non-livestock farming households. Among the Asian studies, eleven focused on dairy farming populations — including cattle owners, dairy farm workers or owners, and livestock keepers — and two on dairy consumers, including rural and non-rural women and health centre attendees. For participant sampling and selection, 27 studies reported random sampling, ten convenience, six purposive, one multistage cluster, and one snowball sampling. Six studies did not specify their sampling approach. Only 30 studies reported participants’ secondary educational attainment and 30 studies participants’ age (not shown). Details of these studies are presented in Table 1.

**Table 1.** Summary characteristics of included studies (n = 51)

**Africa (n = 38)**
| First author (Year)<br>(Reference) | Country | Sample<br>size | Population type | Sampling method | Survey technique | Education (%<br>secondary<br>educated) | ROB (H, M<br>or L) |
| --- | --- | --- | --- | --- | --- | --- | --- |
| Addisu (2020) (28) | Ethiopia | 107 | Cattle owners | Purposive | Questionnaire | 12.1 | M |
| Adesokan (2015)<br>(29) | Nigeria | 196 | Herdsman | Purposive | Interviewer-administered<br>questionnaire | 17.9 | M |
| Almaw (2022) (30) | Ethiopia | 41 | Dairy farm workers<br>or owners | Convenience | Interviewer-administered<br>questionnaire | 21.9 | M |
| Ameni (2007) (31) | Ethiopia | 378 | Cattle owners | Random | Questionnaire | Not specified | L |
| Amenu (2010) (32) | Ethiopia | 98 | Livestock keepers | Purposive | Interviewer-administered<br>questionnaire | 13.3 | L |
| Asebe (2018) (33) | Ethiopia | 384 | Cattle owners | Convenience | Semi-structured<br>questionnaire | 33.3 | L |
| Bihon (2021) (34) | Ethiopia | 349 | Cattle owners | Random | Interviewer-administered<br>questionnaire | 21.8 | L |
| Bonsu (2000) (35) | Ghana | 15 | Cattle owners | Random | Interviewer-administered<br>questionnaire | Not specified | M |
|  |  | 30 | Herdsman | Random | Interviewer-administered<br>questionnaire | Not specified |  |
| Boukary (2011) (36) | Niger | 51 | Cattle owners | Random | Interviewer-administered<br>questionnaire | 0 | M |
| Boukary (2012) (37) | Niger | 1131 | Cattle owners | Random | Interviewer-administered<br>questionnaire | Not specified | H |
| Chileshe (2017) (38) | Zambia | 369 | Dairy farm workers<br>or owners | Not specified | Interviewer-administered<br>questionnaire | Not specified | M |
| Deneke (2022) (39) | Ethiopia | 480 | Dairy farm workers or owners | Random | Questionnaire | Not specified | L |
| Duguma (2017) (40) | Ethiopia | 130 | Pastoralists | Random | Interviewer-administered questionnaire | Not specified | M |
| Gadaga (2016) (41) | Zimbabwe | 102 | Cattle owners | Random | Interviewer-administered questionnaire | 44.1 | L |
| Kang'ethe(2007) (42) | Kenya | 295 | Cattle owners | Not specified | Household questionnaire | Not specified | H |
|  |  | 147 | Consumers | Not specified | Household questionnaire | Not specified |  |
| Katale (2013) (43) | Tanzania | 108 | Pastoralists | Convenience | Interviewer-administered questionnaire | Not specified | H |
| Kazoora (2016) (44) | Uganda | 248 | Dairy farm workers or owners | Multistage cluster | Questionnaires, focus group discussions and interviews | 25.4 | M |
|  |  | 248 | Herdsmen | Multistage cluster | Questionnaires, focus group discussions and interviews | 29.4 |  |
| Kelly(2016) (45) | Cameroon | 46 | Dairy farm workers or owners | Random | Interviewer-administered questionnaire | 10.9 | L |
|  |  | 50 | Pastoralists 1 (Northwest Region) | Random | Interviewer-administered questionnaire | 8.1 |  |
|  |  | 50 | Pastoralists 2 (Vina Division) | Random | Interviewer-administered questionnaire | 2 |  |
| Kemal (2019) (46) | Ethiopia | 43 | Cattle owners | Not specified | Interviewer-administered questionnaire | Not specified | H |
| Kidane (2015) (47) | Ethiopia | 409 | Consumers | Convenience | Questionnaire | 0 | L |
| Kuma (2013) (48) | Ethiopia | 294 | Livestock keepers | Purposive | Interviewer-administered questionnaire | Not specified | L |
| Kusiluka (2006) (49) | Tanzania | 67 | Dairy farm workers or owners | Random | Questionnaire | Not specified | M |
| Lamuka (2018) (50) | Kenya | 75 | Camel pastoralists | Convenience | Questionnaire and focus group discussions | Not specified | H |
| Legesse (2018) (51) | Ethiopia | 460 | Pastoralists | Random | Interviewer-administered questionnaire | 4.2 | L |
| Lencho (2017) (52) | Ethiopia | 100 | Dairy farm workers or owners | Random | Interviewer-administered questionnaire | 24 | L |
| Marange (2020) (53) | South Africa | 110 | Pastoralists | Ballot system | Questionnaire | Not specified | M |
| Meisner (2019) (54) | Uganda | 493 | Pastoralists | Purposive | Interviewer-administered questionnaire | 19.1 | M |
| Mfinanga (2003) (55) | Tanzania | 249 | Consumers | Random | Questionnaire | Not specified | M |
|  |  | 191 | Livestock keepers | Random | Questionnaire | Not specified |  |
| Munyeme (2010) (56) | Zambia | 106 | Cattle owners | Random | Interviewer-administered questionnaire | Not specified | L |
| Obi (2016) (57) | Nigeria | 384 | Herdsmen and dairy farm workers or owners | Convenience | Interviewer-administered questionnaire | 39.1 | L |
| Prudence (2021) (58) | Cameroon | 218 | Herdsmen | Convenience | Interviewer-administered questionnaire | 0 | H |
| Sichewo (2019a) (59) | South Africa | 71 | Cattle owners | Not specified | Questionnaire | 39 | M |
| Sichewo (2019b) (60)* | South Africa | 71 | Cattle owners | Random | Interviewer-administered questionnaire | Not specified | M |
| Tebug (2014) (61) | Malawi | 140 | Dairy farm workers or owners | Random | Questionnaire | 28.6 | M |
| Tora (2022) (62) | Ethiopia | 110 | Cattle owners | Random | Questionnaire | 29.3 | M |
| Tschopp (2013) (63) | Ethiopia | 67 | Dairy farm workers or owners | Random | Questionnaire | Not specified | H |
| Tschopp (2015) (64) | Ethiopia | 130 | Pastoralists | Random | Questionnaire | Not specified | H |
| Tulu (2021) (65) | Ethiopia | 65 | Dairy farm workers or owners | Purposive | Questionnaire | Not specified | H |

\* Follow-up study containing the same sample population as the preceding study by Sichewo et al. (2019a)

Asia (n = 13)
| First author (Year) (Reference) | Country | Sample size | Population type | Sampling method | Survey technique | Education (% secondary educated) | ROB (H, M or L) |
| --- | --- | --- | --- | --- | --- | --- | --- |
| Bagale (2023) (66) | Nepal | 380 | Cattle owners | Random | Interviewer-administered questionnaire | 43.4 | L |
| Hundal (2016) (67) | India | 250 | Dairy farm workers or owners | Random | Interviewer-administered questionnaire | 77.6 | H |
| Islam (2021) (68) | Bangladesh | 404 | Dairy farm workers or owners | Random | Interviewer-administered questionnaire | 30 | L |
| Khattak (2016a) (26) | Pakistan | Not specified | Cattle owners | Random | Interviewer-administered questionnaire | 55 | H |
| Khattak (2016b) (69) | Pakistan | 50 | Livestock keepers | Convenience | Questionnaire | 46 | H |
| Kolhe (2017) (70) | India | 300 | Consumers | Random | Interviewer-administered questionnaire | 29.3 | M |
| Lakra (2020) (27) | India | Not specified | Cattle owners | Snowball | Interviewer-administered questionnaire | Not specified | H |
| Rajkumar (2016) (71) | India | 250 | Livestock keepers | Random | Interviewer-administered questionnaire | 22.8 | M |
| Rajput (2015) (72) | India | 120 | Dairy farm workers or owners | Random | Interviewer-administered questionnaire | Not specified | M |
| Salman (2022) (73) | Iraq | 150 | Consumers | Not specified | Interviewer-administered questionnaire | 42.7 | M |
| Singh (2019) (74) | India | 859 | Livestock keepers | Convenience | Questionnaire | 37.3 | L |
| Ullah (2019) (75) | Pakistan | 190 | Dairy farm workers or owners | Convenience | Interviewer-administered questionnaire | 44.7 | M |
| Ullah (2022) (76) | Pakistan | 200 | Livestock keepers | Random | Interviewer-administered questionnaire | 39.6 | H |

### 3.3 Critical Appraisal of the Studies

Overall, 14 studies were appraised to have a high risk of bias due to insufficient descriptive or statistical analysis, reporting and management of low response rates. Twenty-one studies were deemed to have a medium risk of bias due to incomplete reporting of sampling strategy. Finally, 16 studies were rated with a low risk of bias. All studies were retained for data reporting. The complete critical appraisal is presented in Supplementary Section S.5.

### 3.4 KAP Data Extracted

We extracted 49 KAP fields relevant to bTB. In the following, key themes consistently explored across studies are reported as forest plots. All data fields extracted are presented in Supplementary Section S.3.

### 3.5 Knowledge

Twenty-two knowledge fields under six themes were extracted: knowledge/awareness of tuberculosis or bTB (n=3), knowledge/awareness of zoonotic tuberculosis (n=3), knowledge of tuberculosis signs or symptoms (n=2), knowledge of transmission routes of bTB and zoonoses (n=11), knowledge/awareness of tuberculosis treatment (n=2), and knowledge of bTB mitigation (n=1). We report eight knowledge fields: awareness of tuberculosis, awareness of bTB, awareness that bTB is zoonotic (Figure 2), awareness that raw dairy, raw meat, and inhalation of animal respiratory droplets may result in zoonotic bTB infection (Figure 3), awareness that boiling/pasteurising milk may eliminate pathogens including bTB, and that people can be treated for tuberculosis and possibly cured (Figure 4).

**Figure 2.**
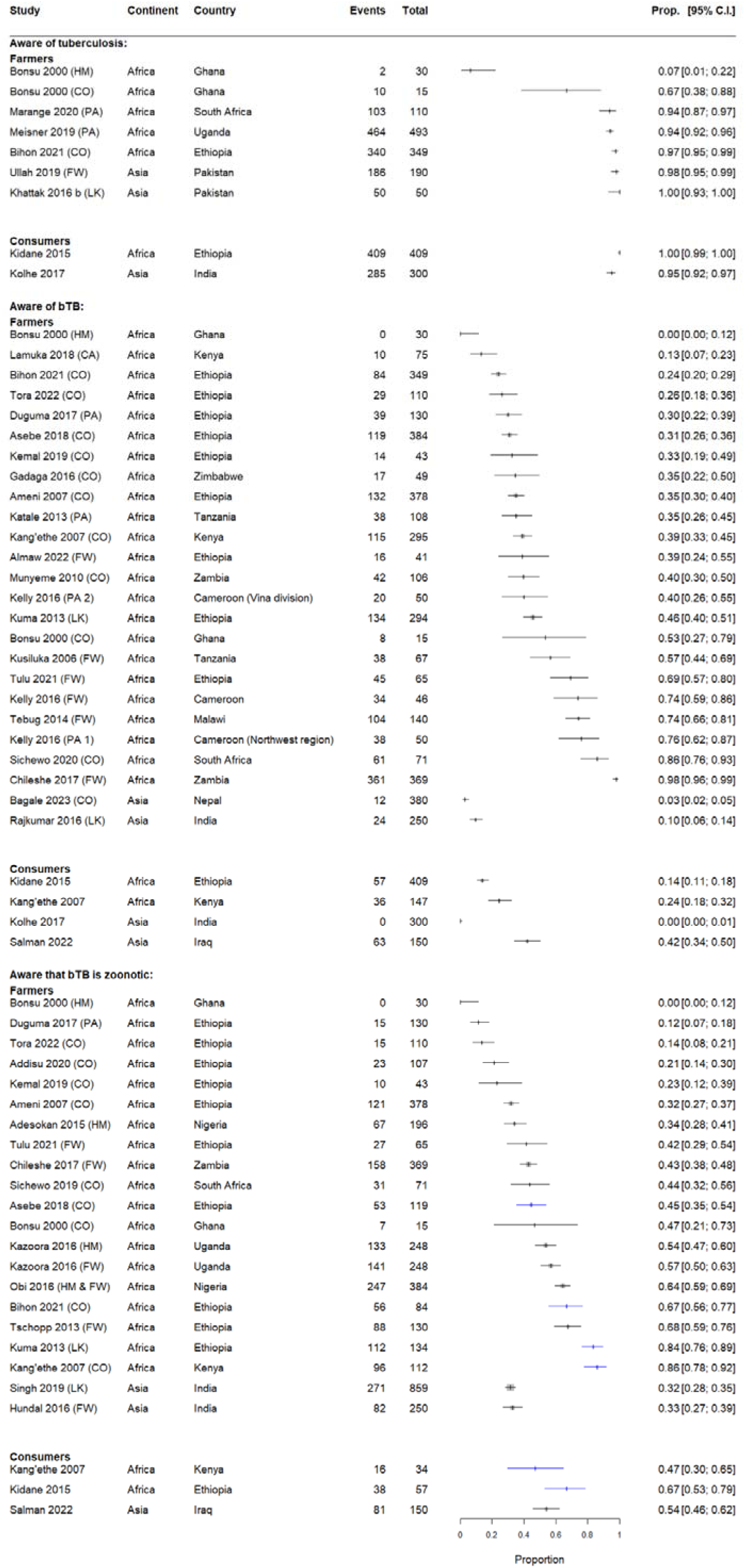
Awareness of tuberculosis, bTB, and the zoonotic potential of bTB amongst dairy farmers and consumers. Boxes represent sample sizes. Blue colouration denotes that the population is a subset of study participants that had prior awareness of bTB. Farmer population types are denoted by; HM (herdsmen), CO (cattle owners), PA (pastoralists), FW (dairy farm workers or owners), LK (livestock keepers), and CA (camel pastoralists). Note that data from one study was excluded from the forest plot due to the lack of reported frequency and total (26).

**Figure 3.**
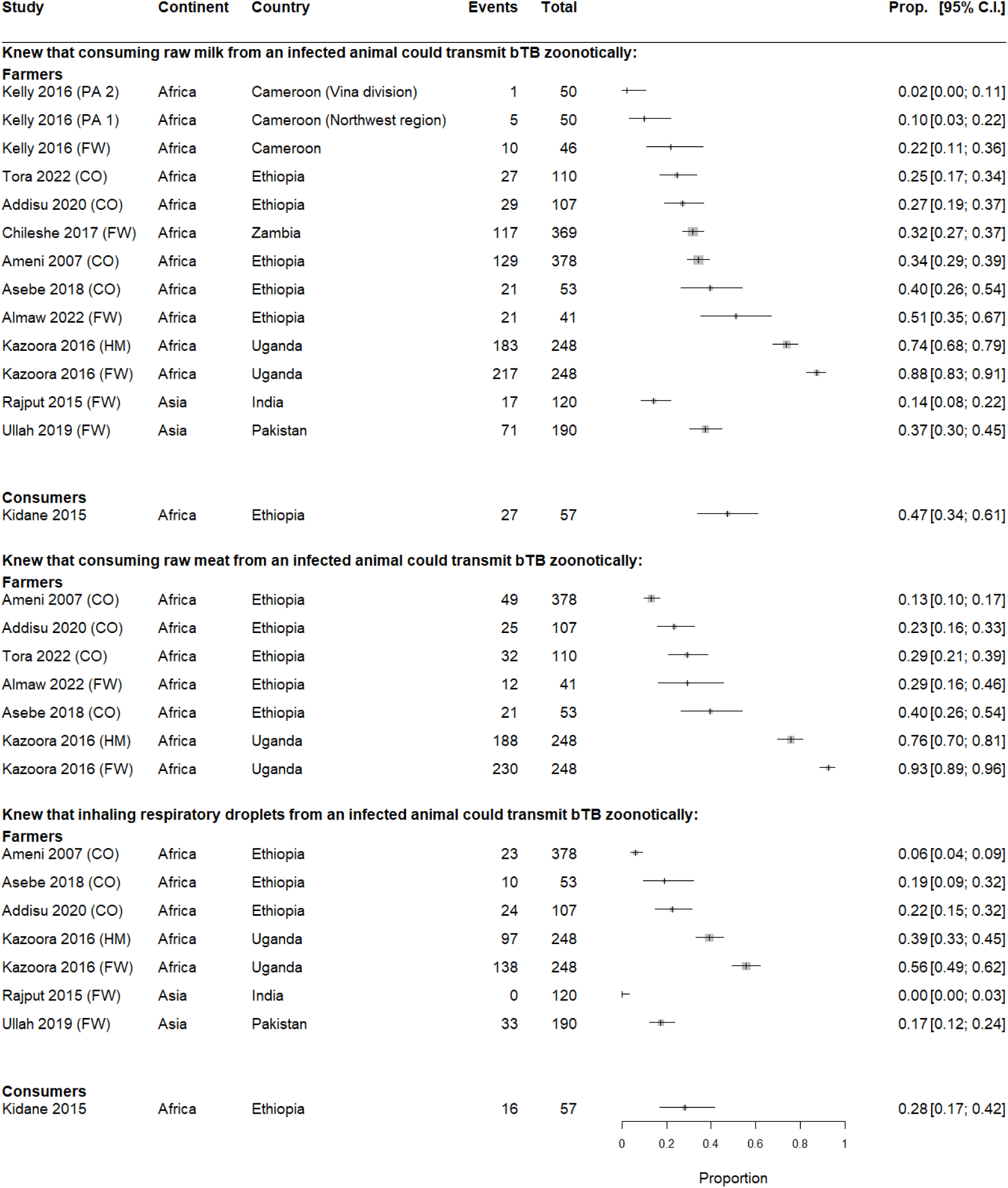
Dairy farmer and consumer knowledge of raw milk, raw meat, and droplet inhalation as potential sources of bTB transmission from animal to human. Boxes represent sample sizes. Farmer population types are denoted by; HM (herdsmen), CO (cattle owners), PA (pastoralists), and FW (dairy farm workers or owners). Note that data from one study was excluded from the forest plot due to the lack of reported frequency and total (26).

**Figure 4.**
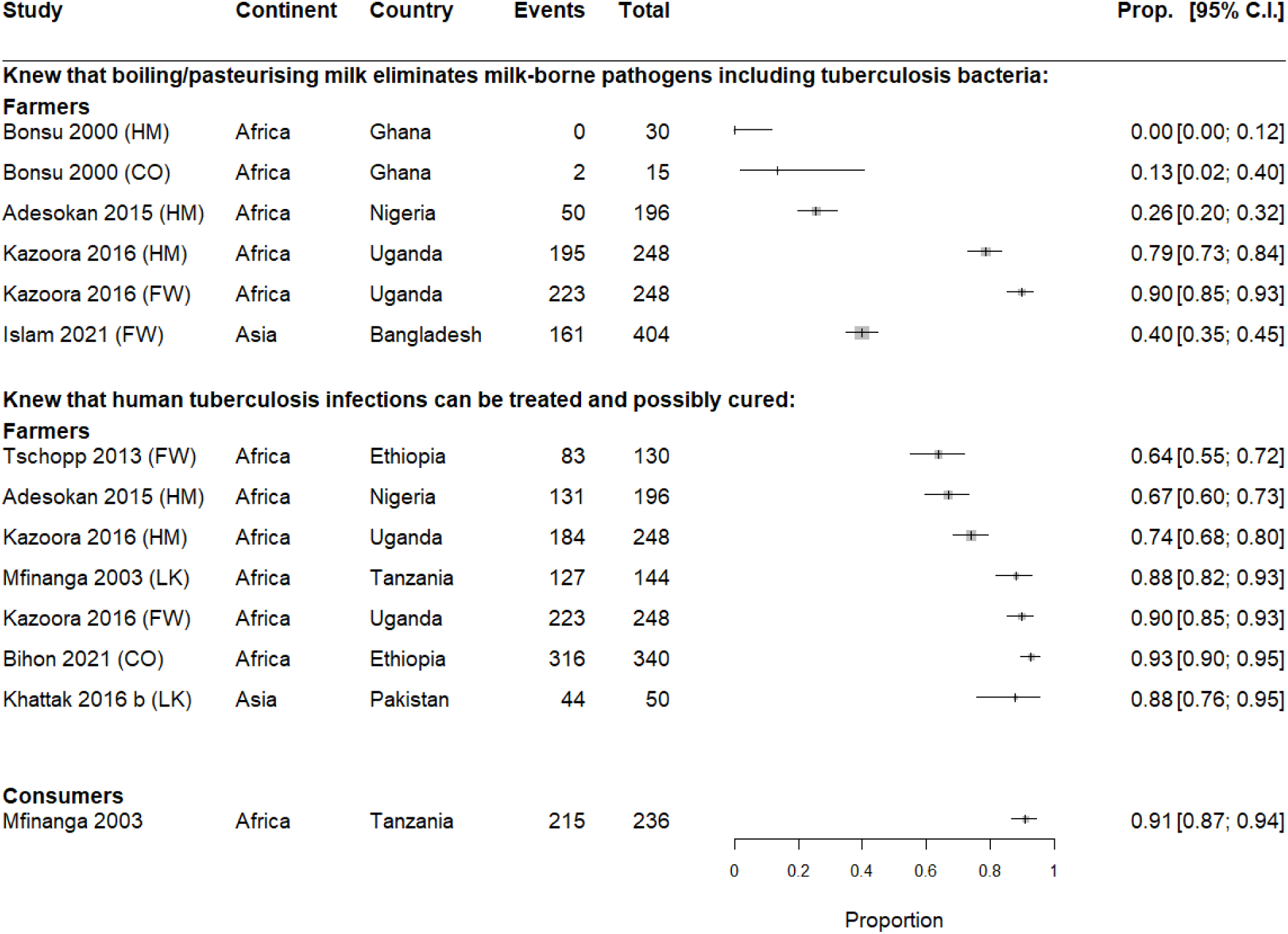
Dairy farmer and consumer knowledge that boiling milk can mitigate bTB transmission, and that human tuberculosis infections can be treated. Boxes represent sample sizes. Farmer population types are denoted by; HM (herdsmen), CO (cattle owners), FW (dairy farm workers or owners), and LK (livestock keepers).

Four African and three Asian studies (one Asian study reported a percentage only and was excluded from the forest plot) quantified tuberculosis awareness among seven farmer populations. Overall, African and Asian farmers investigated were highly aware of tuberculosis, except for one population of herdsmen in Ghana. High levels of tuberculosis awareness among consumers were also reported in Ethiopia and India. For bTB awareness, 23 African and two Asian groups of milk producers were investigated in a total of 22 studies. Collectively, these studies suggest that farmers are less aware of bTB than tuberculosis. Only eight populations had 50% or more participants aware of bTB, and in 12 populations the confidence intervals suggest that bTB was known to less than half of the participants. Six African populations — primarily of farmer workers — had bTB awareness between 50% and 74%, and two over 85%. The Asian studies found almost non-existent bTB awareness among cattle owners and livestock keepers. Studies exploring African and Asian consumers (n=2 each) found low bTB awareness of up to 24% in Africa and 42% in Asia.

Seventeen African and two Asian studies investigated farmers’ awareness that bTB is a zoonosis. Only 12 of these studies explored this zoonotic awareness after asking farmers whether they knew bTB existed (33–35, 38, 40, 42, 46, 48, 62, 65). Of these 12 studies, four reported awareness that bTB is a zoonosis as the proportion of farmers that said ‘yes’ to this question among those who previously confirmed awareness that bTB exists (33, 34, 42, 48) (blue lines, Figure 2). Overall, awareness that bTB is a zoonosis ranged from zero to 86%. There was a high level of zoonotic awareness among farmers who knew bTB exists. Two studies in India suggest low bTB zoonotic awareness in this country. For consumers, all studies exploring awareness that bTB is a zoonosis (n=3) first confirmed consumers’ awareness of bTB, but only two reported it as the proportion of consumers that said ‘yes’ to this question out of those who were aware that bTB exists (42, 47) (blue lines, Figure 2). Overall, consumers showed moderate levels of awareness that bTB is a zoonosis (33, 34, 42, 48).

Few studies investigated whether farmers knew that raw milk and dairy (Africa n=8; Asia n=2) or raw meat (Africa n=6) consumption could result in zoonotic infection with bTB (28, 30, 31, 33, 44, 62). Except for the farmer populations investigated by Kazoora et al. (44), most studies found this knowledge was prevalent among less than 30% of the farmers interviewed. Another six studies that explored all types of milk (46, 51, 52), milk and meat combined (33, 75), all types of meat (46, 76), and one that only reported percentages (26) were excluded from the forest plots but reported similar levels of knowledge well below 50%. Studies investigating farmer awareness of animal respiratory droplets as a source of bTB (Africa n=3; Asia n=1) found limited knowledge among farmers in both continents (Figure 3). Only the Ugandan farmers investigated by Kazoora *et al.* (44) seem to know that boiling/pasteurising milk can eliminate milk-borne pathogens, including bTB. Most of the farmers across the African (n=6) and Asian (n=1) populations knew tuberculosis could be treated and possibly cured (Figure 4).

### 3.6 Attitudes

Eleven attitude fields under four themes were extracted: perceived zoonotic disease transmission pathways (n=4), perceived bTB transmission pathways and mitigation (n=5), attitudes towards treatment (n=1), and milk boiling preference (n=1). None of these attitude fields were explored consistently across studies.

Regardless, attitudes towards transmission pathways of zoonoses and bTB were explored by four studies; three focused on Ethiopian dairy farmers, with the earliest study indicating that less than 25% of participants who had heard of bTB perceived raw meat or dairy consumption to be a bTB transmission pathway (63). Comparatively, two additional Ethiopian studies indicated that raw meat or raw dairy consumption were perceived to be sources of zoonoses by 93% of cattle owners (34, 39). Finally, a study of cattle owners in Zimbabwe found that 8.8% and 47% of participants perceived direct contact with animals and raw milk consumption as risky, and 32% believed that boiling milk reduced the risk of zoonotic diseases (41).

### 3.7 Practices

Fourteen practice fields were extracted, which fall under six themes: milk and dairy consumption (n=7), meat consumption (n=2), animal testing (n=2), biosecurity within the farm, treatment for people, and sharing living space with livestock (n=1 each). Consumption of raw milk/dairy products, consumption of raw meat, and sharing a dwelling with livestock were consistently explored across studies and reported in Figure 5.

**Figure 5.**
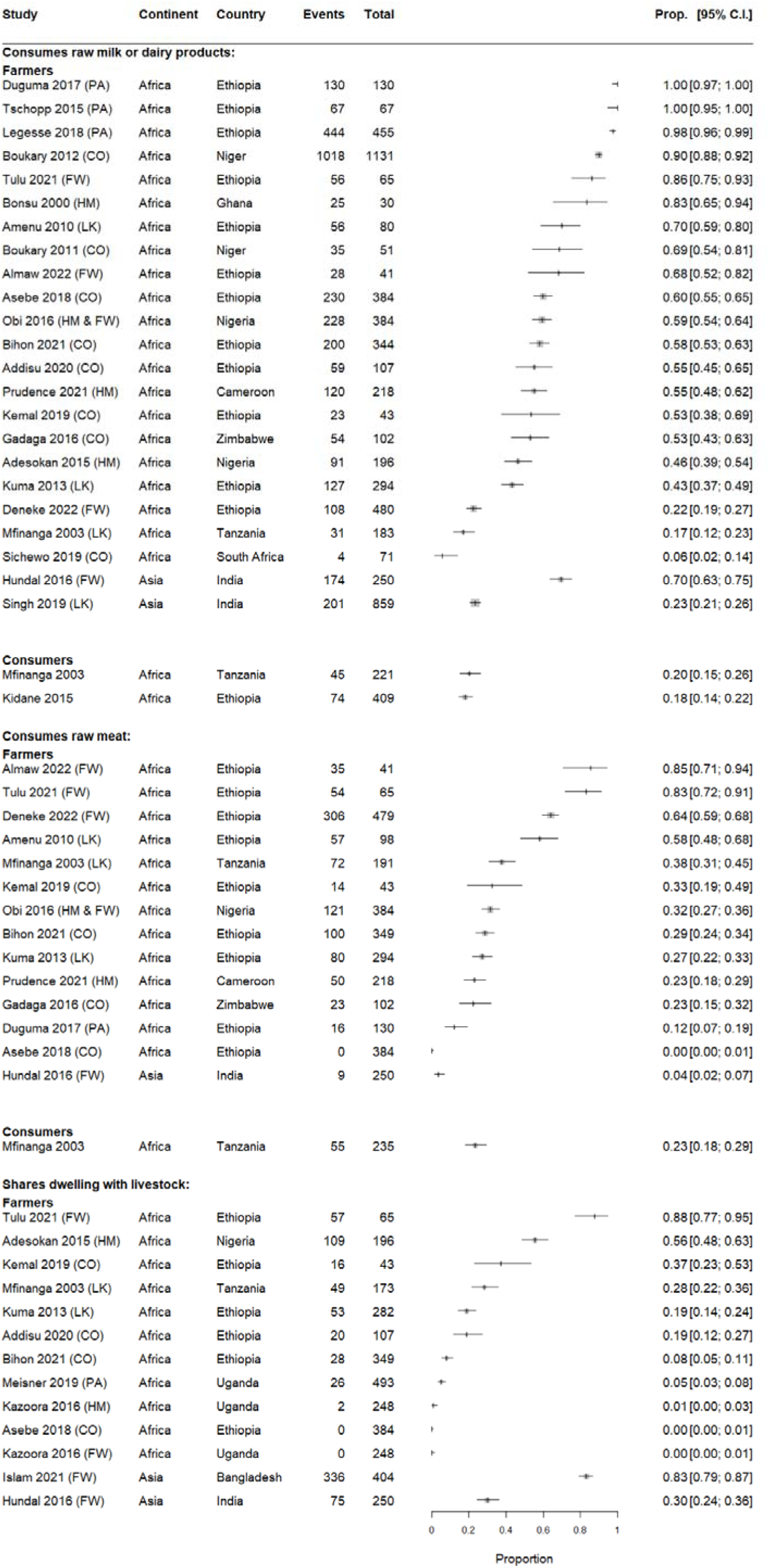
bTB-related practices of dairy farmers and consumers, including raw milk consumption, raw meat consumption, and sharing a dwelling with livestock. Boxes represent sample sizes. Farmer population types are denoted by; HM (herdsmen), CO (cattle owners), PA (pastoralists), FW (dairy farm workers or owners), and LK (livestock keepers). Note that three studies had data excluded from the forest plot: two for not reporting frequency, total or both (27, 52) and one for reporting a frequency larger than the total (62).

Twenty-one African and two Asian studies investigated farmers’ raw milk/dairy consumption. Although a variable practice, half or more participants in most populations and almost all pastoralists in Ethiopia consume raw milk/dairy. Asian studies conducted in India found that 25% of livestock keepers and 75% of farm workers engage in raw milk/dairy consumption. Consumers were only investigated in Africa, who reported seldom consumption of raw milk/dairy.

Dairy farmers in LMICs frequently engage in meat production and consumption. Thirteen studies in Africa and one in Asia investigated farmers’ raw meat consumption. In contrast to raw milk/dairy, in most studies less than half of the farmers reported raw meat consumption. The Asian study found raw meat consumption was uncommon among Indian farm workers (67). A single study that explored raw meat consumption among African consumers reported a low frequency of this practice (55). Finally, African farmers rarely reported sharing a dwelling with their livestock, with less than 20% doing it in eight out of 11 populations studied. In Asia, only 30% of farmers shared a dwelling with livestock in India, while 83% reported doing so in Bangladesh.

## 4 Discussion

In this systematic review, we collated and synthesised data from international studies exploring KAPs towards bTB among dairy producers and consumers in LMICs. The levels of awareness of bTB and its zoonotic potential were surprisingly low despite the same populations having high levels of human tuberculosis awareness. This finding alone illustrates how the long-standing divide between animal and human health messages has obscured the One Health implications of a neglected zoonotic disease that persists among livestock, spreads through food systems, spills into wildlife populations (3, 77) and produces clinically indifferentiable tuberculosis in humans (3). bTB is also naturally resistant to pyrazinamide, undermining the limited arsenal of antimicrobials available to treat tuberculosis (78).

Our review aimed to capture global bTB KAP trends, but only African and South Asian studies met our eligibility criteria. We retrieved several KAP studies conducted in large cattle-producing countries in the Americas, but these targeted non-eligible populations (e.g., veterinarians, medics, and abattoir workers). The final review scope seems to result from three factors: global milk hygiene trends in populations at risk of bTB, and the historical bTB research focus. First, milk pasteurisation was almost universally adopted after the end of World War II across high income countries and LMICs with developed dairy industries, minimising milk-related public health hazards and making bTB a historical public health concern (79).

Second, Africa and Asia are the highest human tuberculosis incidence regions, with almost 2.5 and 4.8 million new cases in 2022, respectively (80, 81); these continents have a high density of milk-producing livestock (82, 83) and neither has robust disease surveillance for bTB (84). Third, over the last 40 years, Africa has been the second most studied bTB region after Europe, and Ethiopia the most studied country globally (85). While our review provides a novel perspective on bTB-related KAPs, global gaps in basic epidemiological research relating to bTB remain, particularly in South and Southeast Asia (9) where the body of bTB research represents only one-fifth of the studies in Africa (85). Our results suggest there is comparatively less bTB knowledge and more risky practices in Asia, yet there are low notification numbers of zoonotic bTB in the region (8) and high bTB prevalence in animal populations (86–88). Similarly, *M. bovis* prevalence is high in cattle herds in the Americas and frequently isolated from dairy products customarily consumed in the region (89–94).

In our review, populations had low awareness of bTB and its zoonotic potential, low awareness of the risks posed by direct contact with animals and consumption of raw animal products, and engaged in several practices that increased their bTB exposure risk. Bovine tuberculosis is intrinsically linked to animals in the food value chain (95), and cattle infected with bTB show low productivity but seldom severe disease or death (96). As milk consumption in Africa and South-East Asia is predicted to grow (97) in several regions with uncontrolled bTB (10), food safety education is likely the cornerstone to realise the public health goals of the zoonotic tuberculosis roadmap in LMICs (98, 99). Our review shows that limited awareness of bTB is accompanied by minimal knowledge of transmission routes and risky practices, a pattern commonly reported for bTB and other zoonoses in various populations, including abattoir and health workers (100–106). Besides these knowledge gaps, we found that attitudes towards bTB were poorly and inconsistently explored across the studies. Beliefs are a critical aspect of risk perception, health-related practice and food safety (18, 107–109); for example, the growing demand for animal protein in Bangladesh incentivised the adulteration of commercial milk, creating population distrust in these products and a belief that raw milk was safer (110). Other examples show how links between beliefs rooted in pre-existing knowledge (or the lack thereof), culture, and experiences result in risky practices. For example, groups consume raw milk because milk boiling generates ‘dead milk’ that lacks nutrients (111) or udder inflammation in the cows producing the milk (112). Other groups seldom consume raw liquid milk but produce dairy products almost exclusively with it (113). Culturally ingrained behaviours can be more resistant to change (114), and require culturally appropriate interventions.

KAP surveys must be designed to avoid or minimise speculative answers. Several studies reporting awareness that bTB is a zoonosis may have used questionnaires that induced participants to report this knowledge, which was not adequately confirmed. Also, studies reported bTB zoonotic awareness either in relation to those who knew bTB previously or everyone irrespective of knowledge, undermining the comparability of results. KAP surveys are relatively simple tools that inform needs, gaps, and locally relevant health intervention priorities when designed with a clear area of enquiry and a well-defined survey population and sampling plan (18). The studies retained for review explored various research questions, used several approaches to explore KAP, and almost half of them used sampling methods that diffused the survey population. Moreover, few studies reported survey validation, how interviewers were recruited and trained, and how the integrity and quality of the data was assured. Unsurprisingly, our risk of bias appraisal suggests that 70% of these studies could have mid- to high risk of bias. In our review, we think the true heterogeneity across studies typically associated with population differences, time of study, and underlying differences in knowledge, attitudes, and practices, may be affected by sampling error and biases attributable to pitfalls in study design (115). While we refrained from producing pooled estimates due to our risk of bias assessment, we are confident the spread of our results closely reflects the true range of variability in knowledge, attitudes, and practices towards bTB across the regions represented.

Our data collation and synthesis had some limitations. First, our synthesis grouped different milk producers — from pastoralists to cattle owners — under the banner of dairy producers. While all these groups make a livelihood from rearing, caring for, and milking their animals, the knowledge, attitudes and practices may significantly vary among groups coexisting in the same areas as shown in our synthesis (35) and elsewhere (116), and therefore should not be seen as equivalent groups. Although we did not find KAP patterns to characterise different milk producers, we made explicit the ‘dairy producer’ group in our results as bespoke interventions may be needed to address their unique needs. We found that studies investigating KAPs of consumers towards bTB are scarce; therefore, we included papers that reported milk and dairy consumption among the general population or subgroups of it, along with any aspects of KAPs towards bTB. Although our synthesis provides insight into milk consumers’ KAPs, targeted research should be warranted as several LMICs lack food safety protocols or integrated food chain surveillance, indicator-based or event-based surveillance for bTB and other zoonoses (117, 118) that put consumers at an unquantified risk of bTB infection. We grouped survey questions across studies under different themes to facilitate KAP synthesis. This thematic grouping reflects the authors’ collective agreement and may be subjective. We acknowledge this by making our extraction tool (Supplementary Section S.2) available to facilitate data appraisal with and without our thematic framing. Finally, we did not consider bTB KAP studies in high-income countries with endemic bTB as most have established animal surveillance systems and milk pasteurisation mandates (10, 119), which translates into KAP studies in these countries focused on farmer’s attitudes towards bTB control, particularly culling of wildlife reservoirs (120–122).

This review addressed an important gap in the bTB literature. Our synthesis of African and Asian studies provides perspective on the risks associated with limited animal bTB surveillance in South-East Asia (10) and the lack of some critical attributes (123) in animal bTB surveillance systems in Africa (118). Livestock production will need to rapidly increase in Africa, Asia and South America to support the accelerated human population growth and associated animal–protein demand in these regions (85) Given this and the increasing importance of alternative livestock, such as camelids, the adaptability of bTB to various species and the isolation of less understood zoonotic *mycobacterium sp.* (124) from dairy animals in South Asia and Africa (8, 124, 125), and reports of anthropozoonosis involving people and dairy cattle elsewhere (126) there is a need for better assessment and monitoring of zoonotic tuberculosis prevalence in food-producing animals. Overall, our review suggests that a “structural One Health” approach (127) that incorporates sociocultural, political, and economic processes and drivers is critical for the efficient, sustainable, and equitable management of bTB (128) to achieve the public health goals of the roadmap for zoonotic tuberculosis. Our review also highlights the need to standardise KAP data collection relevant to bovine tuberculosis to ensure that KAP benchmarks are representative of the population studied, facilitating the monitoring and evaluation of interventions.

## Data Availability

All data produced in the present study are available upon reasonable request to the authors

## Conflict of Interest

The authors have no competing interest to declare.

## Funding

JP Villanueva-Cabezas was supported by The University of Melbourne’s Early Career Research Grant (ECR0662023).

## Notes

### Competing Interest Statement

The authors have declared no competing interest.

### Funding Statement

JP Villanueva-Cabezas received support from The University of Melbourne through an Early Career Research Grant (ECR0662023).

